# Rapid and Portable Quantification of HIV RNA via a Smartphone-enabled Digital CRISPR Device and Deep Learning

**DOI:** 10.1101/2023.05.12.23289911

**Authors:** Hoan T. Ngo, Patarajarin Akarapipad, Pei-Wei Lee, Joon Soo Park, Fan-En Chen, Alexander Y. Trick, Kuangwen Hsieh, Tza-Huei Wang

## Abstract

For the 28.2 million people in the world living with HIV/AIDS and receiving antiretroviral therapy, it is crucial to monitor their HIV viral loads with ease. To this end, rapid and portable diagnostic tools that can quantify HIV RNA are critically needed. We report herein a rapid and quantitative digital CRISPR-assisted HIV RNA detection assay that has been implemented within a portable smartphone-based device as a potential solution. Specifically, we first developed a fluorescence-based reverse transcription recombinase polymerase amplification (RT-RPA)-CRISPR assay for isothermally and rapidly detecting HIV RNA at 42 °C in < 30 min. When realized within a commercial stamp-sized digital chip, this assay yields strongly fluorescent digital reaction wells corresponding to HIV RNA. The isothermal reaction condition and the strong fluorescence in the small digital chip unlock compact thermal and optical components in our device, allowing us to engineer a palm-size (70 × 115 × 80 mm) and lightweight (< 0.6 kg) device. Further leveraging the smartphone, we wrote a custom app to control the device, perform the digital assay, and acquire fluorescence images throughout the assay time. We additionally trained and verified a Deep Learning-based algorithm for analyzing fluorescence images and detecting strongly fluorescent digital reaction wells. Using our smartphone-enabled digital CRISPR device, we were able to detect 75 copies of HIV RNA in 15 min and demonstrate the potential of our device toward convenient monitoring of HIV viral loads and combating the HIV/AIDS epidemic.

## Introduction

With ∼37.9 million people living with HIV/AIDS across the globe, HIV/AIDS remains one of the leading global public health threats (UNAIDS, 2022). Fortunately, revolutionary advances in HIV antiretroviral therapy (ART) in the past two decades have turned this once deadly disease into a manageable chronic condition (Deeks et al., 2013). Nevertheless, for the 28.2 million people living with HIV/AIDS who are receiving ART (UNAIDS, 2022) in the world, access to HIV viral load quantification is essential to monitoring ART efficacy (e.g., suboptimal regimen or resistance to treatment), detecting viral rebound, and preventing onward transmission. As a result, the World Health Organization has been emphasizing the critical yet unmet need for developing diagnostic tools that can improve the accessibility of people living with HIV/AIDS to HIV viral load quantification (WHO, 2016).

Diagnostic tools that can improve the accessibility of people living with HIV/AIDS to HIV viral load quantification should ideally be rapid and portable. Commercial platforms such as GeneXpert or m-PIMA that use RT-PCR to detect HIV are confined to laboratory settings and therefore have limited portability and thus accessibility. In the research arena, existing portable platforms (Drain et al., 2019; Hull et al., 2022; Kadimisetty et al., 2021; Liu et al., 2022; Mauk et al., 2017; Phillips et al., 2019; Trick et al., 2022; Wang et al., 2018), which employ either RT-PCR or RT-LAMP (loop mediated isothermal amplification), generally use bulk-based reactions, which lack accurate quantification and can be slow for detecting low concentrations of targets. These drawbacks call for an alternative for developing rapid, portable, and quantitative diagnostic technologies for HIV viral load quantification.

Compared to bulk-based reactions, digital reactions can detect individual copies of targets and can therefore achieve more accurate quantification. Indeed, there have been reports of digital PCR and digital LAMP for quantitative detection of various pathogens (Rodriguez-Manzano et al., 2016; Xiang et al., 2022; Xu et al., 2021; Yin et al., 2021), including HIV (Shen et al., 2011; Sun et al., 2013). Among digital reactions, digital CRISPR assays have in recent years emerged as an attractive option due to its novelty, absolute quantification, wide dynamic range, and most notably, fast speed (Ding et al., 2021; Luo et al., 2022; Park et al., 2021; Politza et al., 2023; Wu et al., 2021). Despite these advantages, digital CRISPR assays to date have predominantly been performed with bulky instrumentation (e.g., fluorescence microscopy). While smartphones were previously utilized in developing portable digital PCR and digital LAMP devices (Gou et al., 2018; Hu et al., 2020; Liu et al., 2023; Rodriguez-Manzano et al., 2016), the development of smartphone-enabled digital CRISPR devices is in its infancy – the application in HIV viral load quantification has yet to be reported and the speed of the assay has yet to be explored.

In this work, we developed a smartphone-enabled digital CRISPR device for rapid and portable quantification of HIV RNA (Fig. 1). For developing this platform, we first adopted a fluorescence-based reverse transcription recombinase polymerase amplification (RT-RPA)-CRISPR HIV RNA detection assay (Ding et al., 2020) and systematically tuned the assay conditions to accelerate its detection of HIV RNA. We next implemented this accelerated assay within a commercially-available, stamp-sized microfluidic digital chip that we had repurposed for this work. We subsequently engineered a palm-sized but integrated instrument – composed of the smartphone, a fluorescence imaging module, a heating module, and a rechargeable battery – for performing the digital CRISPR assay. We further developed a custom smartphone app for controlling the device with ease, as well as a Deep Learning (DL)-based algorithm for accurately quantifying HIV RNA in the digital chip. Using our smartphone-enabled digital CRISPR device, we were able to detect as low as 75 copies of HIV RNA within 15 min.

**Figure 1.**
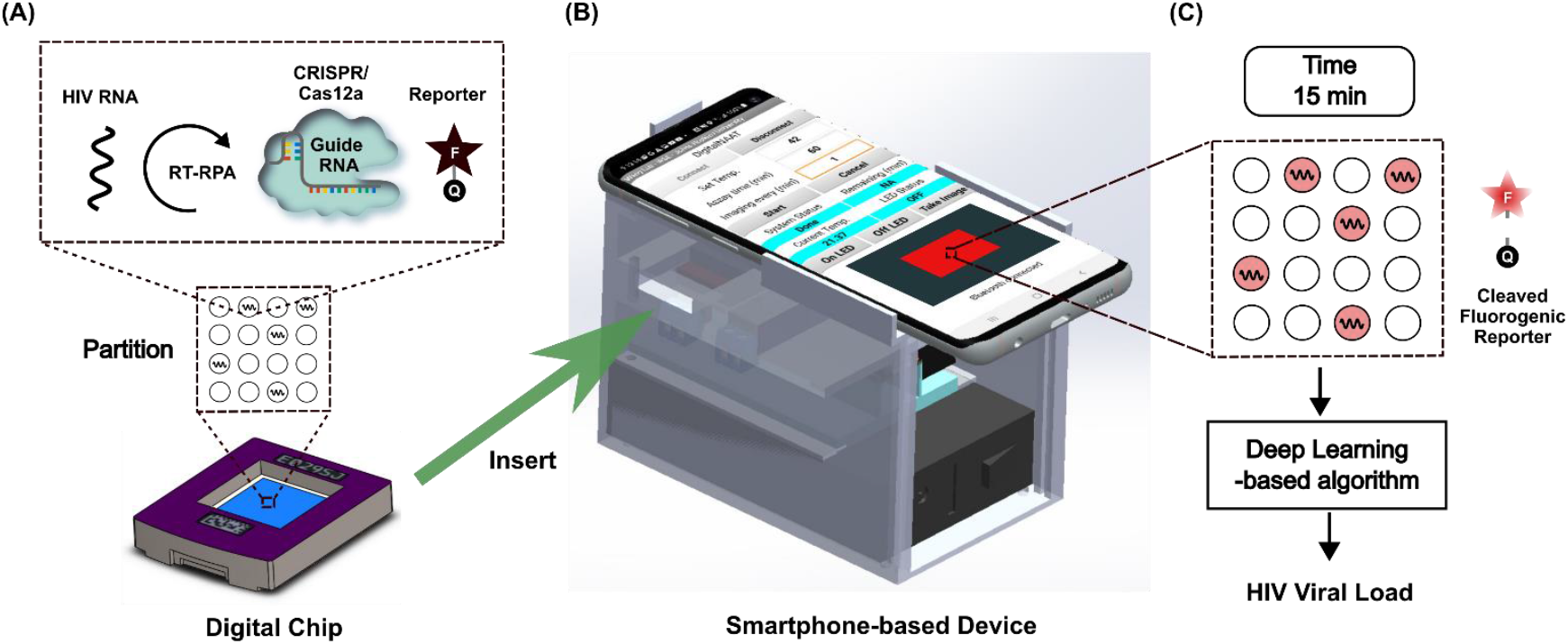
Overview of smartphone-enabled digital CRISPR device for rapid and portable quantification of HIV viral load. Overall workflow: (A) HIV RNA sample and RT-RPA-CRISPR assay reagents are loaded into a commercialonto QuantStudio digital chip.; (B) The digital chip is inserted into thea smartphone-based portable device for HIV RNA target amplification and fluorescence imaging using the smartphone camera.; (C) After 15 min, the fluorescence image is analyzed by a Deep Learning-based algorithm for HIV viral load estimation.

## Results and Discussion

### Overview

The overall workflow is illustrated in Fig. 1. Smartphone-based platform’s components are enclosed within a 3D-printed housing that measures 70 mm × 115 mm × 80 mm (W × L × H) and weights 575 gram. After loading HIV RNA sample and RT-RPA CRISPR assay reagents on a QuantStudio digital chip (Fig. 1A), user insert the chip into the smartphone-based platform and set (1) the desired temperature (e.g., 42 C), (2) the fluorescence imaging frequency (e.g., every 1 min), (3) the total analysis time (e.g., 60 mins) and start the assay using an app installed on smartphone (Fig. 1B). The platform automatically controls temperature for RT-RPA isothermal amplification and captures fluorescence images at the set frequency. The captured images are then analyzed using DL for positive wells detection and HIV viral load quantification (Fig. 1C).

### Rapid CRISPR-assisted HIV RNA detection assay

As a prerequisite for digital CRISPR, we employ published RPA primers and guide RNAs that target HIV gag gene (Ding et al., 2020) and first verify in bulk that the assay can detect HIV RNA (Fig. 2A, Fig. 2B, and Table S1). Here, we focused on improving the speed using benchtop experiments. We used WarmStart RTx reverse transcriptase instead (Chen et al., 2021) (Fig. 2C and Fig. S1). We tuned the reaction temperature and found 42 C outpaced 39 C and 37 C (Fig. 2D and Fig. S2). We also tuned the RTx dose and found 0.75 U/μL can rapidly produce fluorescence signals (Fig. 2E and Fig. S3). As we focused on the speed of the assay, we also tuned the reporter sequence based on previously published reporter sequence and found that TCCCT yields faster signals (Sun et al., 2022) (Fig. 2F and Fig. S4).

**Figure 2.**
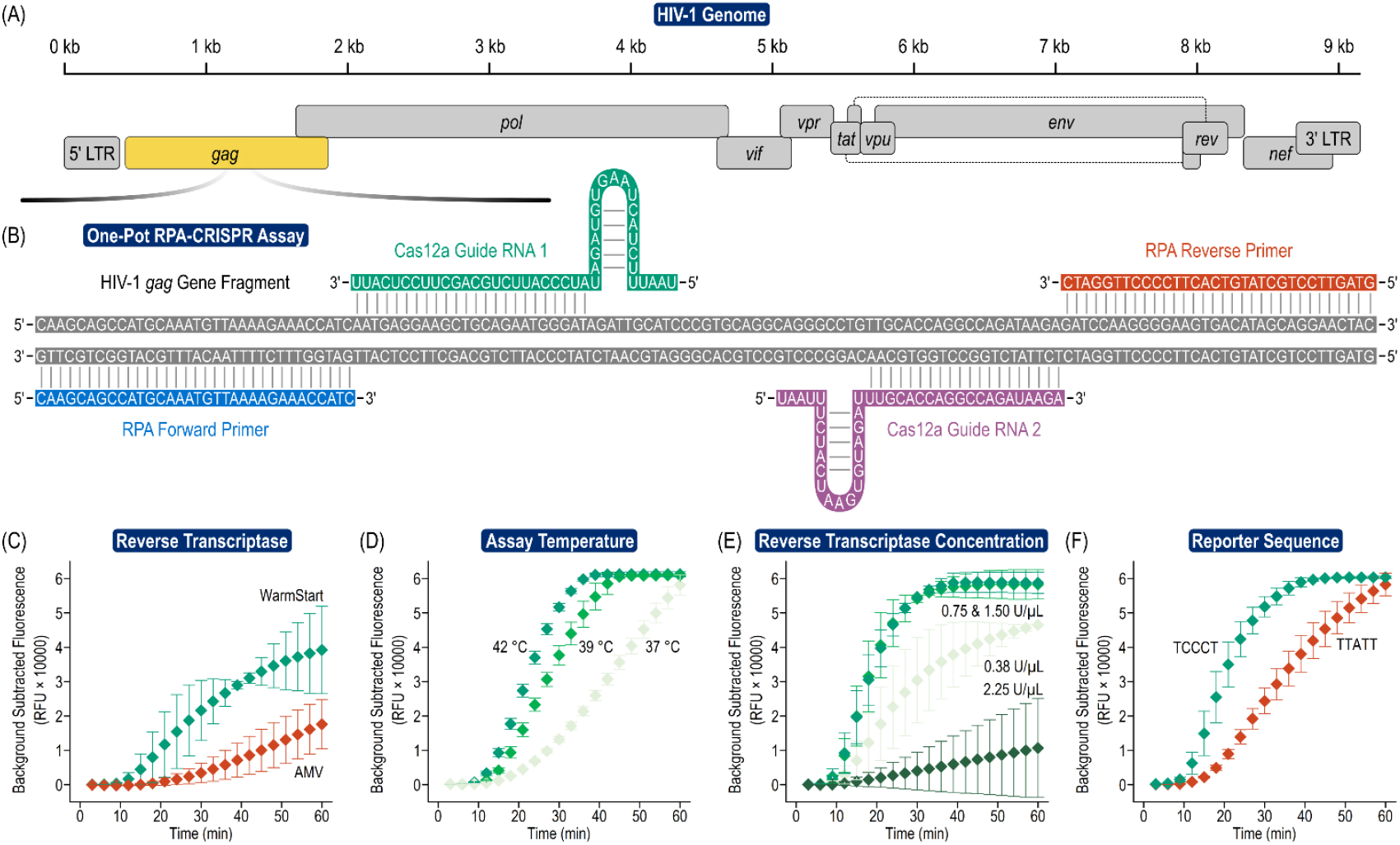
Rapid CRISPR-assisted HIV RNA detection assay. (A) The assay targets HIV gag gene and (B) features a pair of RPA primers and a pair of guide RNA. By tuning (C) reverse transcriptase, (D) assay temperature, (E) reverse transcriptase concentration, and (F) reporter sequence, the assay can detect 1,000 HIV RNA copies with plateauing fluorescence signals within 30 min.

### Compact digital assay platform

We engineered an integrated instrument that can perform digital CRISPR-assisted assay. The instrument is composed of a smartphone, a “fluorescence macrophotography” module, a heater, and control circuitry (Fig. 3A). The instrument is built around commercial QuantStudio digital chips – commercially available digital chips with 750-pL wells positioned in a 20,000-well array – as use of a commercially available digital chip can help improve accessibility when compared to custom microfluidic chips. We selected a smartphone (Samsung Galaxy S10) as the testbed for our instrument because this phone is relatively inexpensive yet has a solid camera with 12 megapixel and the capacity for manual focus adjustments.

**Figure 3.**
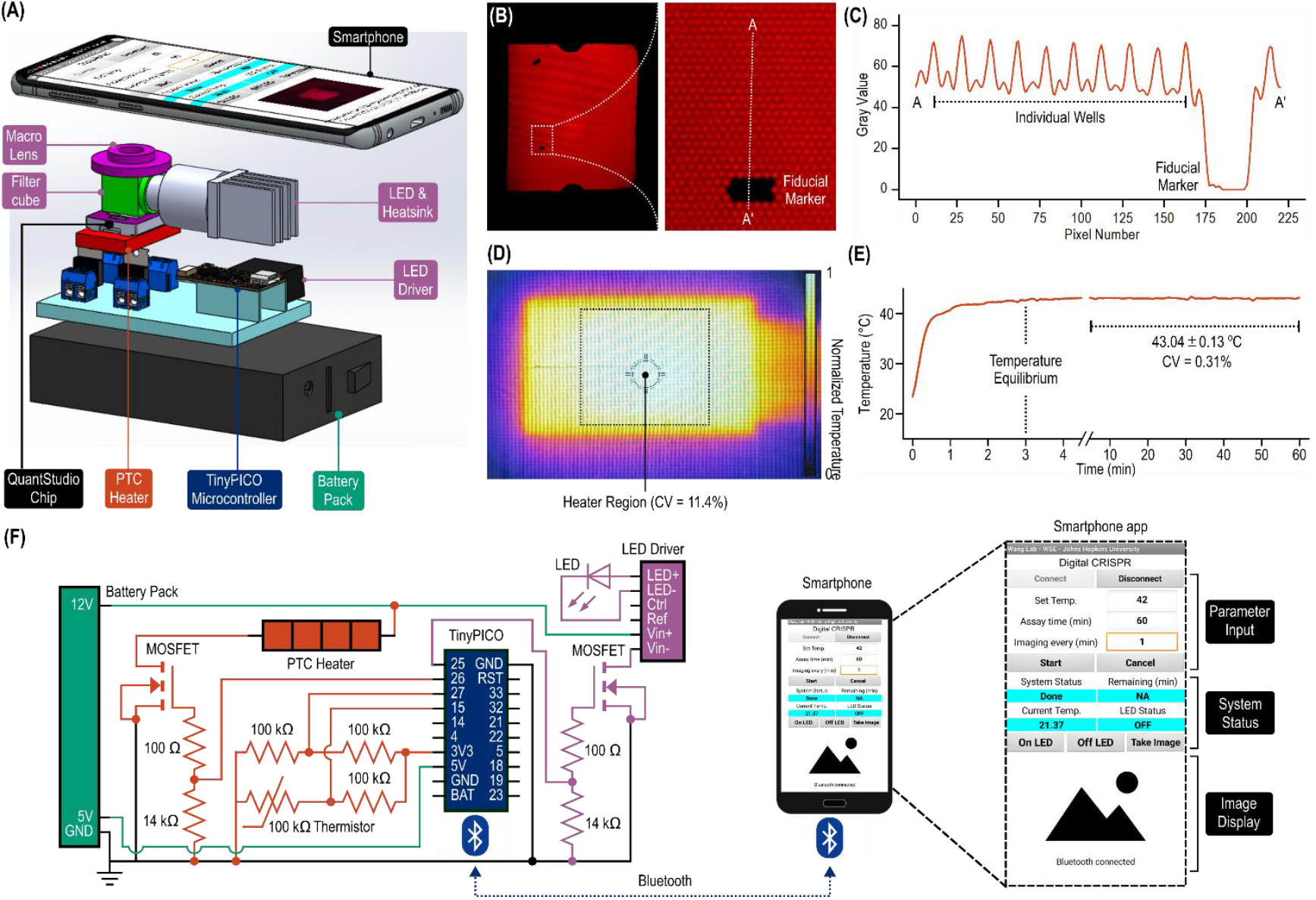
Portable digital CRISPR device. (A) In addition to the smartphone, the key components of the device include a macro lens, a LED (and its accompanying heat sink), a custom fluorescence filter cube, a LED driver, a positive temperature coefficient (PTC) ceramic heater, control circuitry featuring a TinyPICO microcomputer, and a battery pack. These components are integrated within a 3D printed housing (not shown) such that the macro lens positioned beneath and aligned to the smartphone to view the QuantStudio digital chip. (B) The fluorescence macrophotography module – composed of the macro lens, the custom fluorescence cube, and the LED – enables the smartphone to view the entire QuantStudio chip while resolving individual digital wells. (C) The fluorescence intensities in the wells are stronger from the pitch between adjacent wells. (D) The compact PTC ceramic heater delivers uniform temperature across the heater region that is comparable to the size of the QuantStudio chip. (E) Once initiated, the PTC ceramic heater can reach temperature equilibrium in ∼3 min and maintain stable temperature throughout the course of the reaction. (F) The control circuitry includes a TinyPICO microcomputer, a circuit for measuring and controlling temperature of the PTC ceramic heater, and a circuit for turning the LED on and off. A custom smartphone app is developed to input necessary parameters, display system status, and show fluorescence images throughout the assay.

As an alternative to assembling sophisticated optics in our platform, we developed a simple fluorescence macrophotography module – composed of a macro lens, a LED, and a custom filter cube – that could image the QuantStudio chip with sufficient resolution and field of view. For putting together the fluorescence macrophotography module, we first identified a solid macro lens with sufficient resolution and field of view for imaging the QuantStudio chip. To do so, we purchased 6 commercial smartphone-compatible macro lenses and used them to image a standard resolution target and a standard distortion target at their maximum and minimum working distances (Fig. S5), from which we determined their magnification, field of view, resolution, distortion, and range of working distance (Fig. S6). We note a tradeoff between magnification and working distance during our evaluation – because we designed our fluorescence macrophotography module to incorporate a custom miniature fluorescence filter cube, we needed to disqualify a macro lens with excellent magnification but insufficient working distance for fitting the filter cube which is 17.5 mm height. Upon our evaluation, we selected a macro lens that – combined with the camera lens – achieved a 35.9 line pairs/mm resolution (13.92 μm line width), 0.99% pincushion distortion, and 22 mm by 16.5 mm field of view at 19 mm working distance (Fig. S7 and S8). To complete the fluorescence macrophotography module, we assembled an excitation filter, a dichroic mirror, an emission filter, and a compact LED via a 3D printed holder. We subsequently evaluated the fully assembled fluorescence macrophotography module by detecting a QuantStudio chip loaded with 10 μM Cy5 fluorescence dye, a concentration that is comparable to the reporter concentration in the eventual RPA-CRISPR assay. The addition of the fluorescence macrophotography module allowed us to use the smartphone to image the entire chip while resolving individual digital wells (Fig. 3B & 3C). As a sanity check, we attempted to image the same QuantStudio chip without the fluorescence macrophotography module and failed to acquire useful images for analysis (data not shown).

For isothermal temperature control, we employed a positive temperature coefficient (PTC) ceramic heater (Fig. S9) (Ngo et al., 2022), which has several advantages including fast warm-up, relatively uniform heating, and self-regulating, therefore being safe and energy efficient. Indeed, using the PTC ceramic heater, we achieved relatively uniform temperature across the heater surface (Fig. 3D). The warm-up time was fast, reaching the set temperature from room temperature in ∼3 min and staying relatively stable after that (Fig. 3E).

All components of the platform (PTC heater, excitation LED, and smartphone’s camera) are controlled by an electronics circuit and a custom smartphone app. The electronics circuit was composed of a TinyPICO ESP32 microcontroller, a circuit for measuring and controlling temperature of the PTC ceramic heater, and a circuit for controlling the LED (Fig. 3F). The smartphone app allows user set up the assay temperature, assay time, and fluorescence imaging frequency. After user set up the above parameters and press start, the app sends these parameters to the TinyPICO ESP32 microcontroller via Bluetooth. Upon receiving parameters, the TinyPICO ESP32 microcontroller measure and control temperature of the PTC heater to match the set temperature using a PID controller. Once the set temperature is reached, the TinyPICO ESP32 microcontroller turn on LED for fluorescence excitation and inform the app to capture fluorescence images at the set imaging frequency. Flowcharts of the TinyPICO ESP32 microcontroller’s program and the smartphone app are illustrated in Fig. S10 and S11.

### Facile identification of positive digital wells

Using the integrated instrument (Fig. S12), we first demonstrated digital CRISPR-assisted detection of HIV RNA in QuantStudio chip. Here, when we input 7,500 copies of HIV RNA and performed the reaction for 60 min, we observed strongly fluorescent wells (i.e., positive), indicating successful amplification and detection of HIV RNA (Fig. 4A).

**Figure 4.**
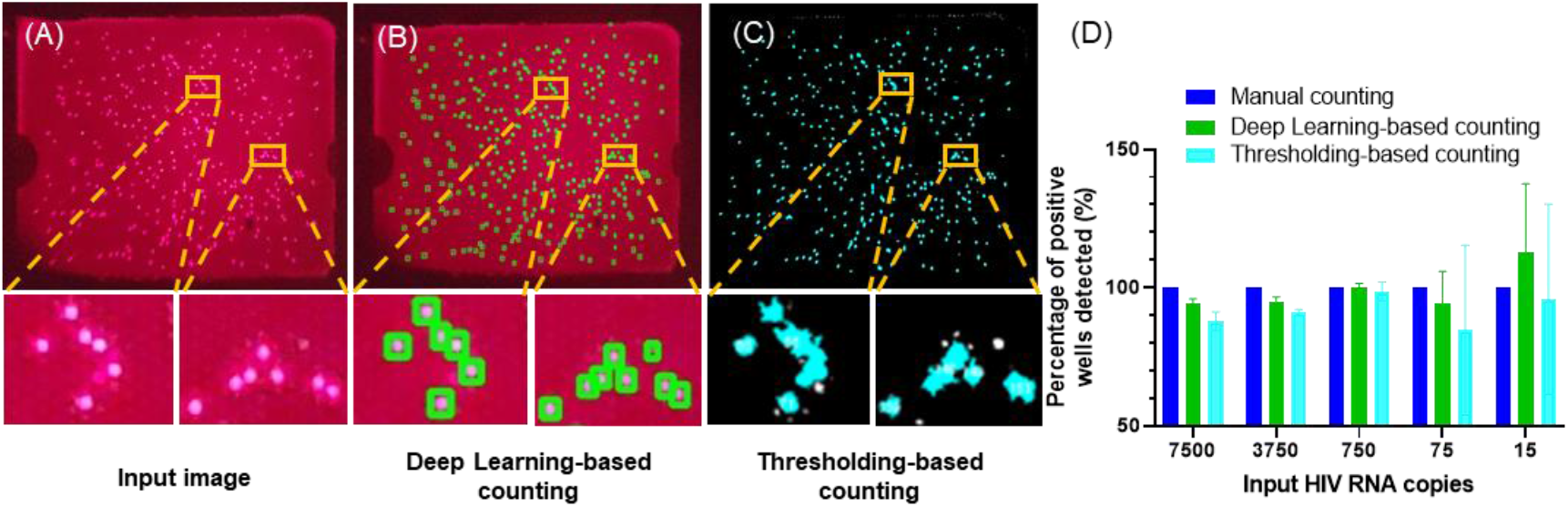
Positive well detection using Deep Learning and Thresholding. (A) raw QuantStudio image; (B) Positive wells detected by DL; (C) Positive wells detected by thresholding; (D) Percentage of positive wells detected by DL and thresholding (normalized to manual counting) at various input HIV RNA copy numbers. Error bars represent standard deviations (n = 3).

We explored two methods for quantifying positive wells in fluorescence images of QuantStudio chip – a DL-based workflow and a conventional thresholding-based workflow. For the DL workflow, since the size of positive wells in the images is relatively small, to detect them, we used a YOLOv5 object detection architecture in combination with an image tiling approach (Fig. S13). Image tiling approach has been shown to be effective in detecting small objects in images (Unel et al., 2019). Upon DL model training, DL can reliably detect 333 positive wells (Fig. 4B) compared to 348 positive wells detected by manual counting (Fig. S14). Notably, DL can detect positive wells that are close to each other. For thresholding-based workflow, we used auto thresholding and particle analysis functions in ImageJ to detect the positive wells. Thresholding can also adequately detect positive wells, though we noticed that it had difficulty in detecting positive wells that are close to each other. During thresholding, these wells are often merged, and the number of positive wells detected decreased to 310 (Fig. 4C). Using manual counting as a reference method, DL method outstripped thresholding-based method in detecting positive wells across different input HIV RNA copy numbers (Fig. 4D). It is noteworthy that at low input HIV RNA copy number (15 copies and 75 copies), error of both methods increased. The reason is that at these low input HIV RNA copies, the number of positive wells is relatively low (a few to ten positive wells). Therefore, just one counting error by DL or thresholding will be equivalent to 10%-30% error (or more) in comparison to manual counting.

### Rapid and quantitative detection of HIV RNA

In addition to end-point detection, our instrument also supports real-time monitoring of the digital CRISPR assay and provides means to accelerate the assay time. Indeed, for the sample of 7,500 HIV RNA copies, 234 positive wells could be observed as short as 15 min (Fig. 5A). As time increases, the number of positive wells increased but began to plateau after 30 min (Fig. 5A). In other words, the number of positive wells remained almost the same after 30 min. To verify this, we quantified the percentage of positive wells normalized to the number of positive wells detected at 60 min for different input HIV RNA copy numbers. Based on this method, we found that 39% of positive wells were detectable by 10 min, 71% of positive wells were detectable by 15 min, and nearly all positive wells (93%) could be detected by 30 min (Fig. 5B). These results indicate that the assay time could be shortened to 15 min while retaining satisfactory capability in quantifying HIV RNA.

**Figure 5.**
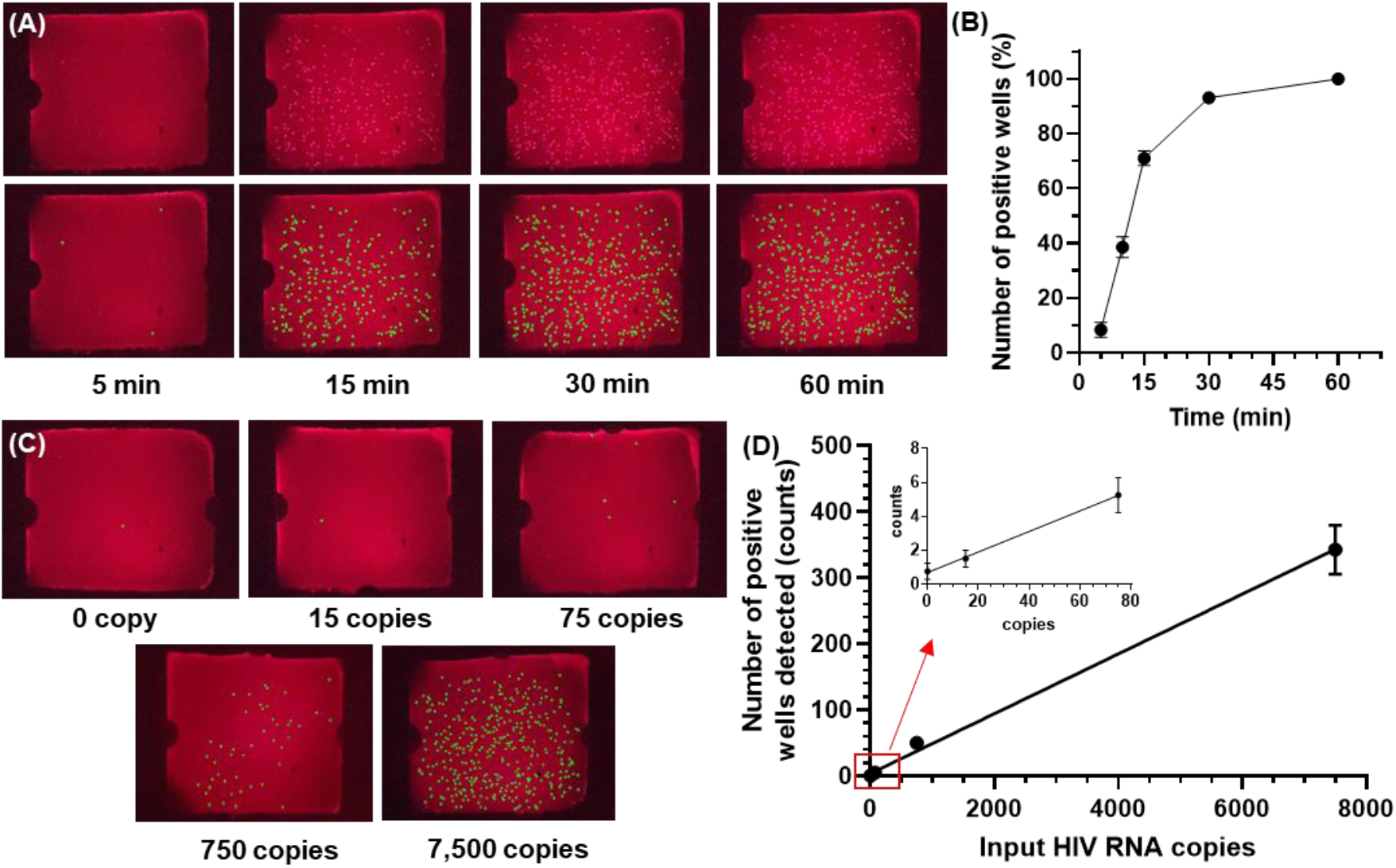
Digital CRISPR-assisted HIV RNA detection. (A) raw and DL-analyzed QuantStudio chip images of 500 HIV RNA cp/μL input target concentration at 5 min, 15 min, 30 min, and 60 min; (B) Percentage of positive wells normalized to the number of positive wells detected at 60 min (n = 8); (C) DL-analyzed QuantStudio images of different target concentrations 0, 15, 75, 750, and 7,500 HIV RNA copies at 15 min; (D) Dose response curve at the 15 min time (n = 3). Error bars represent standard error of the mean.

Finally, we demonstrated sensitive detection of HIV RNA in 15 min using our instrument. Here, we tested 0, 15, 75, 750, and 7,500 input HIV RNA copy number. As expected, as the input copy number decreased, we observed fewer positive wells (Fig. 5C). The standard curve is linear across 2 orders of magnitude. As few as 75 HIV RNA can be detected (Fig. 5D).

## Discussion

The rapidity, portability, and quantitation capacity of our smartphone-enabled digital CRISPR device were achieved by synergistically integrating various components. First, digitization of the CRISPR assay not only enabled quantification but also accelerated the assay speed, especially for low concentrations of RNA. Such an improvement was possible because digitization of each RNA molecule within a digital well created a uniformly high local concentration across the entire digital chip, and as a result the assay speed became independent of the RNA concentration. Second, to date, smartphones have been combined with lenses into microscopes, which have high resolution but small field of view, or have been directly used without any modification (i.e., no external lens) as imagers, which have large field of view but low resolution. Fluorescence macrophotography presented herein represents a simple strategy for achieving both high resolution and large field of view, this allowing our smartphone to image the entire QuantStudio digital chip while retaining single-well resolution. Using our compact fluorescence macrophotography module for the detection of our digital CRISPR reaction was possible also because the reaction produced strong fluorescence signals within positive digital wells. Indeed, nearly all positive wells that were detected by a sensitive but bulky fluorescence microscope could also be detected by our compact fluorescence macrophotography module (Fig. S15). Finally, the low-temperature isothermal reaction condition of our digital CRISPR reaction alleviated the need for a bulky power source for heating. Taking advantage of such synergy of the components was key to developing our smartphone-enabled device. In fact, despite its significantly lower cost and smaller footprint, our smartphone-enabled device showed comparable performance against benchtop instruments (Fig. S16).

To our knowledge, our smartphone-enabled digital CRISPR device represents the first such device for rapid and portable quantification of HIV RNA. Despite the advance, we see several areas for improvement. First, as our device currently only detects HIV RNA, sample preparation (e.g., HIV RNA extraction) still needs to be incorporated. To this end, nucleic acid extraction based on magnetic beads (Fan et al., 2023; Jiang et al., 2023; Kang et al., 2021; Ngo et al., 2023; Pearlman et al., 2020; Trick et al., 2022), cellulose paper (Zou et al., 2017), or FTA membrane (Kadimisetty et al., 2021; Ndunguru et al., 2005) offer possible solutions. It may also be feasible to adopt a custom digital chip with integrated nucleic acid extraction capability (Yin et al., 2020). Another area for improvement is assay sensitivity. The current sensitivity of 75 copies requires >= 375 μL of blood to meet the virological failure threshold of 200 copies/mL. Toward improving the assay sensitivity, our digital RPA-CRISPR assay may be further adjusted (e.g., tuning the Cas12a/crRNA concentration (Shao et al., 2022) or modified (e.g., incorporating photoactivation of Cas12a (Hu et al., 2022)). Alternatively, incorporating a sample concentration step as part of the sample preparation process prior to digital CRISPR may be an option. Third, real-time reporting of HIV viral load directly on the smartphone can improve user-friendliness, as our device currently completes a digital CRISPR reaction before the fluorescence images are exported and analyzed. Finally, the current device costs ∼700 US dollars without the smartphone, which is significantly cheaper than commercial or research platforms but still too expensive as an accessible diagnostic tool. Thus, cost reduction by employing more cost-effective optical components represents an important objective. These improvements can elevate the prospect of our smartphone-enabled digital CRISPR device as a practical tool for HIV viral load monitoring.

## Conclusions

We report herein a smartphone-enabled digital CRISPR device for rapid and portable quantification of HIV RNA. For the development of our device, we first tuned an isothermal (42 °C) RT-RPA-CRISPR assay to accelerate its detection of HIV RNA to < 30 min. We then digitized this assay using commercially available QuantStudio chips, which allowed us to achieve digital quantification without the need of more complicated microfluidic devices and associated instrumentation. We subsequently employed compact optical components, thermal components, and microcontroller to engineer an integrated device that is palm-sized (70 × 115 × 80 mm), lightweight (< 0.6 kg), and battery-powered. We additionally developed a custom smartphone app to control the device and perform the digital assay, which simplified device operation while also leveraging the full capability of the smartphone. We also developed a DL-based algorithm for analyzing fluorescence images and detecting strongly fluorescent digital reaction wells. Using our device, we were able to qualitatively detect HIV RNA in 10 min and accurately quantify HIV RNA in 15 min, across a dynamic range of 3 orders of magnitude and down to a LOD of 75 copies. We note that the development of our smartphone-enabled digital CRISPR device represents a synergistic integration of the RT-RPA-CRISPR assay, QuantStudio chip, optical and thermal components, and the smartphone. Areas for improvement for the current device include incorporation of sample preparation, detection of viral particles, ease-of-use, and reduction of cost. With promising current results and avenues for future improvement, our smartphone-enabled digital CRISPR device may emerge as a useful solution toward convenient monitoring of HIV viral loads for the 28.2 million people in the world living with HIV/AIDS and receiving ART and toward combating HIV/AIDS at large.

## Methods

### Benchtop RPA-CRISPR assay

All oligonucleotides, including RPA primers, Cas12a-guide RNAs, Alexa647-labeled single-stranded DNA (ssDNA) fluorogenic reporter were purchased from Integrated DNA Technologies (IDT; Coralville, IA). Both Cas12a-guide RNAs were modified with IDT’s proprietary 5’ AltR1 and 3’ AltR2 modifications. Lyophilized Cas12a-guide RNA was reconstituted in DEPC-treated water (Thermo Fisher Scientific, Waltham, MA) at 20 μM. Lyophilized DNA primers, DNA reporter, and DNA probe were reconstituted in nuclease-free water (Promega, Madison, WI) at 100 μM. EnGen Lba (Lachnospiraceae bacterium ND2006) Cas12a (Cpf1) (M0653; 100 μM), WarmStart® RTx reverse transcriptase (M0380; 15000 U/ml), Avian Myeloblastosis Virus (AMV) reverse transcriptase (M0277; 10000 U/mL), and bovine serum albumin (BSA; B9000; 20 mg/mL) were purchased from New England BioLabs (Ipswich, MA). TwistAmp Basic kits were purchased from TwistDx Limited (Maidenhead, United Kingdom). All reconstituted oligonucleotides and aforementioned reagents were stored at -20 °C.

To prepare a 10-μL reaction mixture for CRISPR-Cas12a-assisted RT-RPA, we used 1× rehydrated TwistAmp Basic Reaction mix, 0.32 μM of each RPA primer, 0.64 μM of each Cas12a-guide RNA, 4 μM of Alexa647-labeled ssDNA fluorogenic reporter, 0.64 μM of EnGen Lba Cas12a, 0.75 U/μL of WarmStart RTx reverse transcriptase, 0.01 mg/mL of BSA, and 14 mM of MgOAc, unless stated otherwise (e.g., in Fig. S1 through S4). The assembly was performed in a PCR hood (AirClean Systems, Creedmoor, NC). First, each dried pellet of TwistAmp Basic Reaction mix was resuspended in 29.5 μL of rehydration buffer to prepare 1.7× rehydrated TwistAmp Basic Reaction mix. Next, a master mix of all components except WarmStart RTx reverse transcriptase, BSA, and MgOAc was prepared in a 1.5 mL protein low-binding microcentrifuge tube (MilliporeSigma, Burlington, MA) and incubated at room temperature for 10 min to allow the formation of Cas12a-guide RNA complexes. Then, WarmStart RTx reverse transcriptase and BSA were added to the master mix. The master mix was transferred to a biosafety cabinet (The Baker Company, Sanford, ME) to prevent contamination. Inside the biosafety cabinet, the master mix was divided into 8-μL MgOAc-free reaction mixture aliquots and supplemented with 1 μL of 140 mM MgOAc and 1 μL of 1000 copies/μL of synthetic HIV RNA to obtain the final 10-μL CRISPR-Cas12a-assisted RT-RPA reaction mixture.

### Brightfield Characterization of macro lens

To derive the resolution of lenses, the resolution target from Edmund (USAF 1951 Glass Slide Resolution Targets, AZ) was used, and each lens was attached to the smartphone camera and set about 3 cm away from the target to take an image. The image was analyzed using ImageJ by drawing a line across each set of the 3-line-pairs and deriving the line profile plot using Analyze option in ImageJ. Then, to consider if the line pair can be resolved, the mean and standard deviation of the maximum peaks (white lines) were calculated, and according to empirical rule, assumed normal distribution, Mean ± 3.Std covers 99.7% of data, so if the minimum peaks (represented black lines) fall within 99.7% range of the white lines, it means that they couldn’t be distinguished from the white lines completely; therefore, the resolution of the lens will be based on the line pair one set before this one.

The distortion percentage of the lenses was evaluated using a Distortion Target (Edmund Optics, AZ) and calculated by relating the distance of the center to the corner of the distortion target image as the image actual corner (AC) indicated by blue arrow and the distance of the center to the center edge corner (CC) determined by drawing straight lines from center of two edges to form a corner indicated by red arrow as shown in Fig. S8 and Equation 1. If the points in the FOV appeared too close to the center, the distortion is negative (barrel distortion). On the other hand, if the points are too far away from the center, the distortion is positive (pincushion distortion).

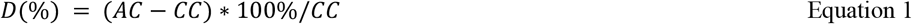

### Construction of the phone-based platform

Temperature was measured by a thermistor (GA100K6MCD1, TE Connectivity) epoxied to a PTC heater (50W AC DC 12V, Bolsen Tech). A red LED (627nm Rebel LED on a SinkPAD-II 20mm Star Base - 53 lm @ 350mA) for fluorescence excitation was controlled by a 350mA LED driver (BuckPuck 3021-D-E-350, LEDdynamics Inc.). Power supplied to the PTC heater and the LED driver was controlled by two N-CH MOSFETs (IRLB8721PBF, Infineon), which were in turn controlled by an ESP-32 microcontroller (TinyPICO, Unexpected Maker). Macro lens (25X, MACTREM) was used for focusing smartphone camera on the QuantStudio chip. Filter cube was comprised of an excitation bandpass filter 624nm CWL - 40nm bandwidth, a dichroic filter 660nm, and an emission bandpass filter 692nm CWL - 40nm bandwidth (Edmund Optics). The whole platform was powered by a 12V rechargeable Lithium ion battery 3000mAh (TalentCell). User set temperature, imaging frequency, and total analysis time using an app (developed using MIT App Inventor 2, http://ai2.appinventor.mit.edu/#4872056792809472) on a smartphone (Samsung galaxy S10 phone). These parameters were sent via Bluetooth to the ESP-32 microcontroller which will control the temperature of the PTC heater, turn on LED for fluorescence excitation, and inform the smartphone to capture fluorescence images. For the app to access the smartphone camera, we used the Taifun Camera extension (Version 6 as of 2020-07-13 14:00. Extension Version: 6, Date Built: 2020-07-13, More information http://puravidaapps.com/camera.php). Using this extension, we set the smartphone camera in continuous-picture focus mode, auto white balance, and minimal exposure compensation. To achieve the best focus images, before experiments with test samples, the smartphone camera focus was calibrated using a QuantStudio chip loaded with Cy5 fluorescence dyes. In brief, after inserting the chip into the platform and turning on the LED, user imaged the chip using Pro mode of the built-in Camera app of Samsung Galaxy S10 phone (ISO 400, 1/15 F1.5, white balance: A 4500K, manually focus until the image is sharpest). After the smartphone camera focus calibration step, the platform is ready for use.

### Detection of HIV RNA using the phone-based platform

To test the platform, a master mix containing RPA buffer 1X, HIV forward and reverse primer each 0.32 μM, LbCas12a 0.64 μM, HIV crRNA1 and crRNA2 each 0.64 μM, Alexa647-labeled fluorogenic reporter 4 μM, WarmStart reverse transcriptase 0.75 U/μL, BSA 0.01mg/mL, Tween-20 0.1%, and DEPC water was prepared. After that, master mix aliquots (each 13.5 μL) were spiked with 0.75 μL of synthetic HIV RNA sequences containing 0, 15, 75, 750, or 7,500 copies and 0.75 μL of 280 mM MgOAC. The samples were loaded into QuantStudio chips using QuantStudio™ 3D Digital PCR Chip Loader, followed by inserting QuantStudio chips into the platform for RT-RPA amplification at 42 ºC for 60 min and fluorescence imaging every 1 min.

### Image analysis using Deep Learning

First, we trained an object detection deep learning model based on YoLoV5 ([Ultralytics](https://github.com/ultralytics/yolov5)) for detecting positive wells in fluorescence image of QuantStudio chip. Since the size of positive well objects in Quantstudio chip fluorescence image is relatively small, to detect them we employed an image tiling approach during the deep learning model training and deployment (Fig. S13). Specifically, several fluorescence images (4032 pixel x 3024 pixel) were cut into 416 pixel x 416 pixel patches, resulting into 143 tile images of 416 pixel x 416 pixel size. Positive wells in the 143 tile images were labeled using a labeling tool. After labeling, the 143 tile images were divided into a training set (97 tile images), a validation set (26 tile images), and a testing set (20 tile images). Data augmentation (e.g., flip, 90 degree rotate, crop, brightness, exposure, blur…) was applied to the training set (maximum version size 3X), creating a final set of 97*3 = 291 tile images for training. No data augmentation was applied to validation set and testing set. Training was done using Google Colab with 150 epochs (completed in 0.127 hours) and resulted in mAP50 of 0.991 and mAP50-95 of 0.546. For deployment, weights of the trained model were exported and saved to a personal computer and a Python code was developed for (1) loading fluorescence image of interest, (2) estimating chip region from the loaded image, and (3) detecting positive wells in the estimated chip region using the trained deep learning model and image tiling (Fig. S13). Image analysis time was less than 1 min/image.

## Data Availability

All data produced in the present work are contained in the manuscript

## Declaration of competing interest

The authors declare that they have no known competing financial interests or personal relationships that could have appeared to influence the work reported in this paper.

## Acknowledgments

This work was supported by funding through the National Institutes of Health (R01AI138978, R01AI137272, and R61AI154628). K.H. is grateful for a developmental grant from the Johns Hopkins University Center for AIDS Research, a National Institutes of Health funded program (P30AI094189).

## Notes

### Competing Interest Statement

The authors have declared no competing interest.

