## Supporting Information document for "Rapid and Portable Quantification of HIV RNA via a Smartphone-enabled Digital CRISPR Device and Deep Learning"

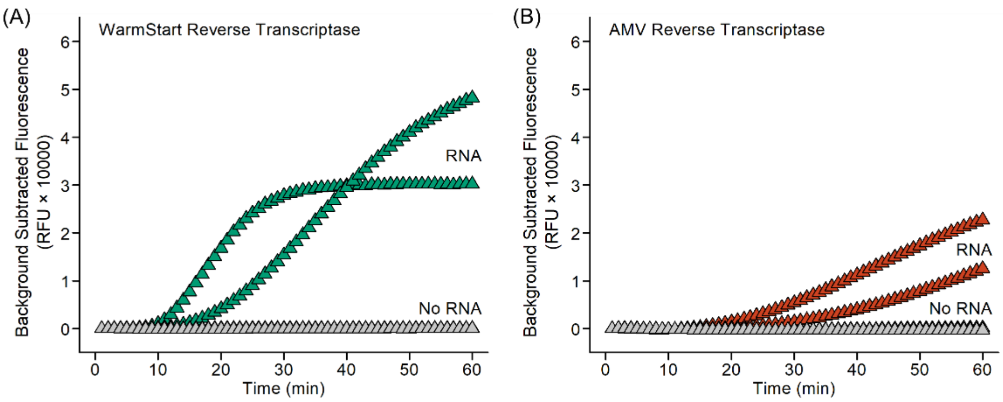

**Figure S1. Reverse transcriptase comparison.** Benchtop real-time amplification curves of RT-RPA-CRISPR assays with (A) WarmStart reverse transcriptase and (B) AMV reverse transcriptase show that WarmStart reverse transcriptase outstrips AMV reverse transcriptase. Here, each 10-µL RT-RPA-CRISPR reaction mixture consists of 1× rehydrated TwistAmp Basic Reaction mix, 0.32 µM of each RPA primer, 0.64 µM of each Cas12a-guide RNA, 4 µM Alexa647-labeled ssDNA fluorogenic reporter (TTATT), 0.64 µM EnGen Lba Cas12a, 0.75 U/µL of either WarmStart RTx reverse transcriptase or AMV reverse transcriptase, 0.01 mg/mL BSA, 14 mM MgOAc, and either 1000 copies or 0 copy of HIV RNA. The reactions are performed in a Bio-Rad CFX96 Touch Real-Time PCR Detection System at 42 °C for 60 min, and the fluorescence signals are measured every 1 min. The fluorescence signals measured by the Bio-Rad CFX96 system are first exported without baseline subtraction (*i.e.*, under "No Baseline Subtraction" mode in the CFX Manager Software). Then, using Microsoft Excel, the fluorescence signals from every reaction are background subtracted (i.e., $F - F_{t=1}$). Finally, all plots are plotted using Origin. Results in Figures S2 – S4 are exported, background subtracted, and plotted in the same manner as Figure S1.

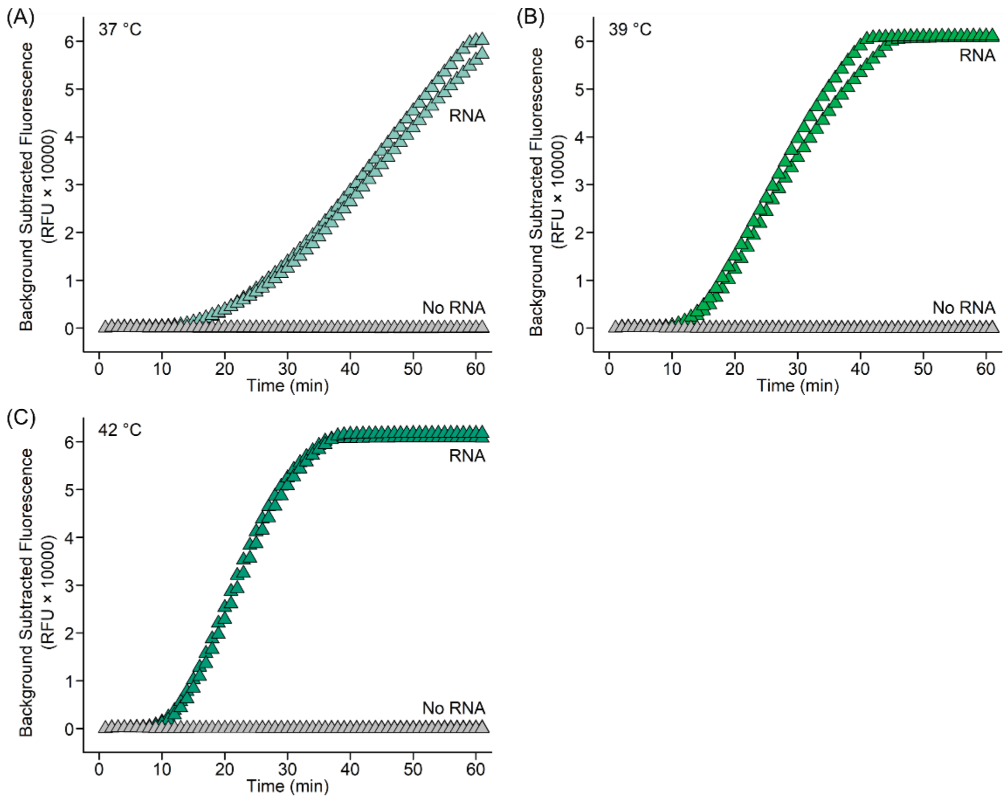

**Figure S2. Assay temperature comparison.** Benchtop real-time amplification curves of RT-RPA-CRISPR assays performed at (A) 37 °C, (B) 39 °C, and (C) 42 °C reveal that 42 °C enables the fastest reaction. Here, each 10-µL RT-RPA-CRISPR reaction mixture consists of 1× rehydrated TwistAmp Basic Reaction mix, 0.32 µM of each RPA primer, 0.64 µM of each Cas12a-guide RNA, 4 µM Alexa647-labeled ssDNA fluorogenic reporter (TTATT), 0.64 µM EnGen Lba Cas12a, 0.75 U/µL WarmStart RTx reverse transcriptase, 0.01 mg/mL BSA, 14 mM MgOAc, and either 1000 copies or 0 copy of HIV RNA. The reactions are performed in a Bio-Rad CFX96 Touch Real-Time PCR Detection System at 37 °C, 39 °C, and 42 °C using its built-in gradient function for 60 min, and the fluorescence signals are measured every 1 min.

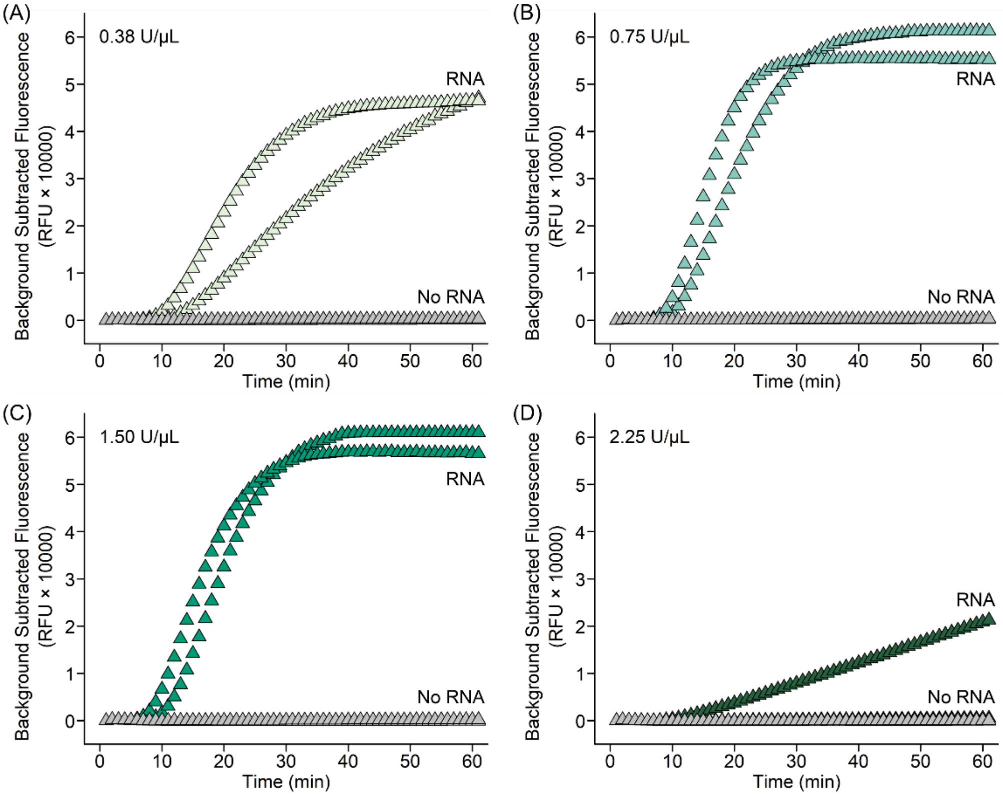

**Figure S3. Reverse transcriptase concentration comparison.** Benchtop real-time amplification curves of RT-RPA-CRISPR assays with (A) 0.38 U/µL, (B) 0.75 U/µL, (C) 1.50 U/µL, and (D) 2.25 U/µL reverse transcriptase reveal that 0.75 U/µL and 1.50 U/µL have comparably rapid reaction kinetics, whereas 2.25 U/µL yields poor amplification, suggesting that excessive reverse transcriptase can in fact cause amplification failure. Here, each 10-µL RT-RPA-CRISPR reaction mixture consists of 1× rehydrated TwistAmp Basic Reaction mix, 0.32 µM of each RPA primer, 0.64 µM of each Cas12a-guide RNA, 4 µM Alexa647-labeled ssDNA fluorogenic reporter (TTATT), 0.64 µM EnGen Lba Cas12a, either 0.38 U/µL, 0.75 U/µL, 1.50 U/µL, or 2.25 U/µL WarmStart RTx reverse transcriptase, 0.01 mg/mL BSA, 14 mM MgOAc, and either 1000 copies or 0 copy of HIV RNA. The reactions are performed in a Bio-Rad CFX96 Touch Real-Time PCR Detection System at 42 °C for 60 min, and the fluorescence signals are measured every 1 min. .

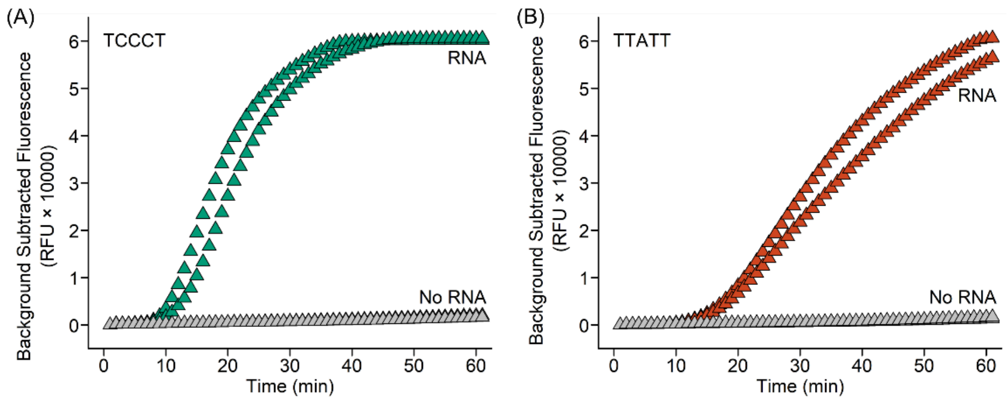

**Figure S4. Fluorogenic reporter comparison.** Benchtop real-time amplification curves of RT-RPA-CRISPR assays with (A) TCCCT ssDNA reporter and (B) TTATT ssDNA reporter reveal that TCCCT ssDNA reporter outperforms TTATT ssDNA reporter. Here, each 10-µL RT-RPA-CRISPR reaction mixture consists of 1× rehydrated TwistAmp Basic Reaction mix, 0.32 µM of each RPA primer, 0.64 µM of each Cas12a-guide RNA, 4 µM Alexa647-labeled ssDNA fluorogenic reporter (either TCCCT or TTATT), 0.64 µM EnGen Lba Cas12a, 0.75 U/µL WarmStart RTx reverse transcriptase, 0.01 mg/mL BSA, 14 mM MgOAc, and either 1000 copies or 0 copy of HIV RNA. The reactions are performed in a Bio-Rad CFX96 Touch Real-Time PCR Detection System at 42 °C for 60 min, and the fluorescence signals are measured every 1 min.

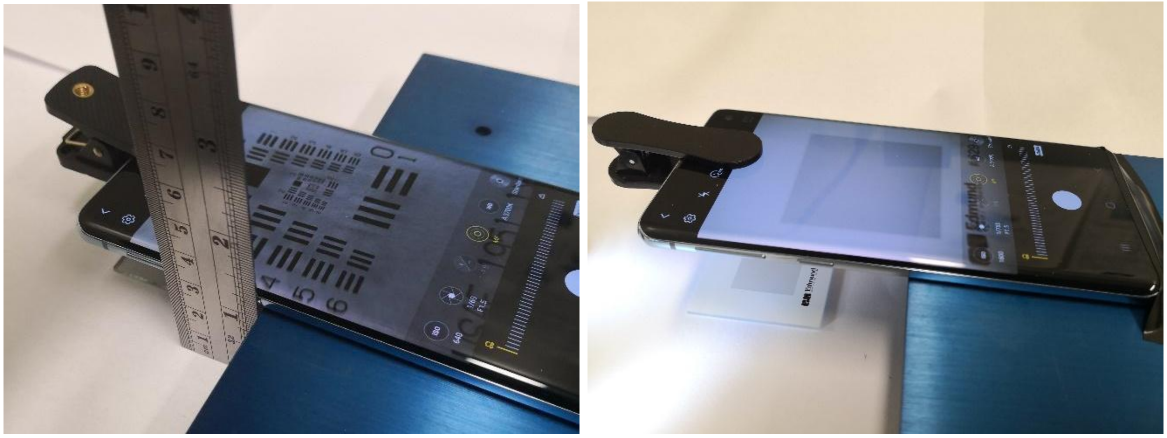

**Figure S5. Setup for characterizing macro lenses in brightfield.** Each macro lens is attached to the testbed smartphone, placed the assembly on an adjustable pedestal, and manually focused the camera onto (left) a resolution target (2 in. × 2 in. USAF 1951 1× resolution target, stock #38-257, Edmund Optics) and (right) a distortion target (2 in. × 2 in. 0.0625 mm dot, low reflect grid distortion target, stock #62-949, Edmund Optics). For both targets, images were taken at the minimum working distance, as measured by a ruler, when the images appeared clearly focused.

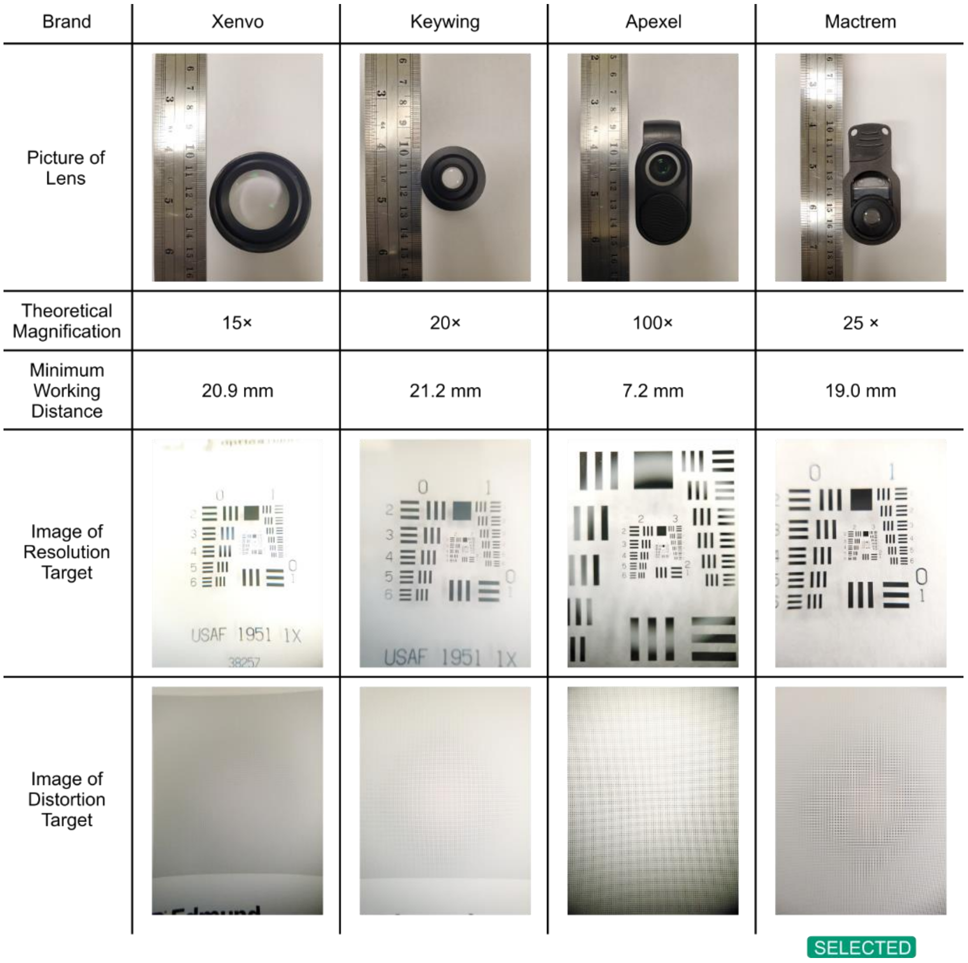

**Figure S6. Evaluation of four macro lenses.** Minimum working distances, images of the resolution target, and images of the distortion target of the four macro lenses were compared toward selecting the appropriate macro lens for the fluorescence macrophotography module. The Xenvo macro lens had insufficient magnification. The Apexel macro lens is in fact marketed as a microscope for smartphones and indeed had excellent magnification. Unfortunately, its small working distance was insufficient for fitting the custom fluorescence filter cube. Both the Keywing macro lens and the Mactrem macro lens had sufficient magnifications and working distances, but the Mactrem macro lens was selected due to its greater magnification.

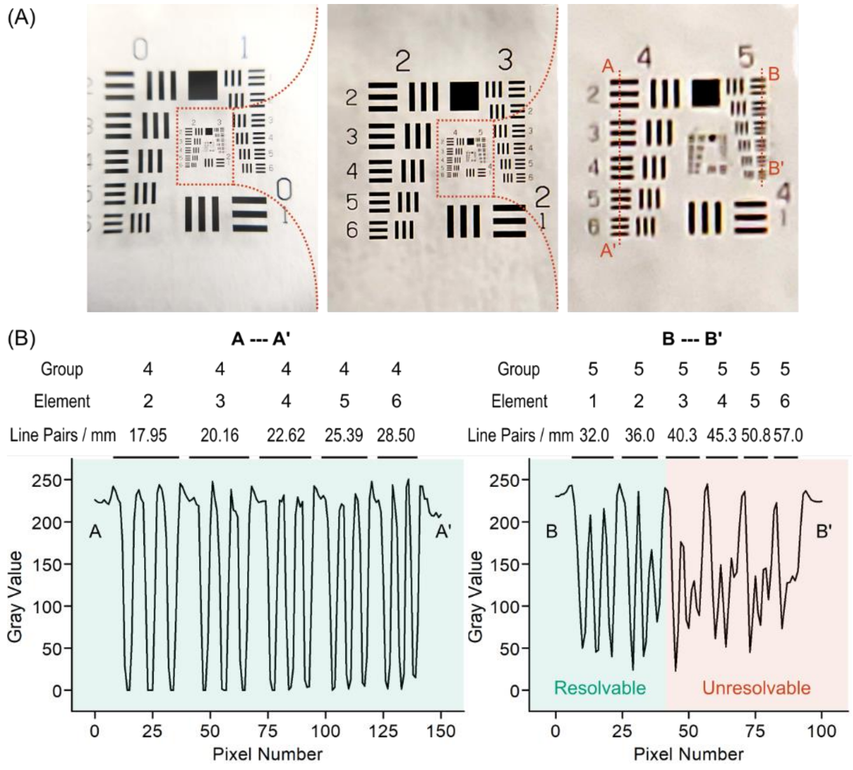

**Figure S7. Quantification of resolution of Mactrem macro lens.** (A) The image of the resolution target taken by the Mactrem macro lens was analyzed using ImageJ by drawing lines across elements (i.e., sets of three line-pairs) in group 4 (A – A') and group 5 (B – B') of the resolution target and subsequently deriving (B) line profile plots using the Analyze function in ImageJ. To determine if an element is resolvable, the mean and the standard deviation (SD) of the three white lines from the element – corresponding to the three peaks in the line profile plots – were first calculated and then used to set a threshold at mean – 3SD. For each element, if its threshold remains above all three corresponding valleys in the line profile plots – corresponding to the black lines from the element – then element is considered resolvable. Based on such calculation, group 5 element 2, which represents 36.0 line pairs per mm (equivalent to ~13.9 µm line and ~13.9 µm pitch), is resolvable. This resolution is sufficient for resolving the 60-µm-wide digital wells in the QuantStudio digital chip.

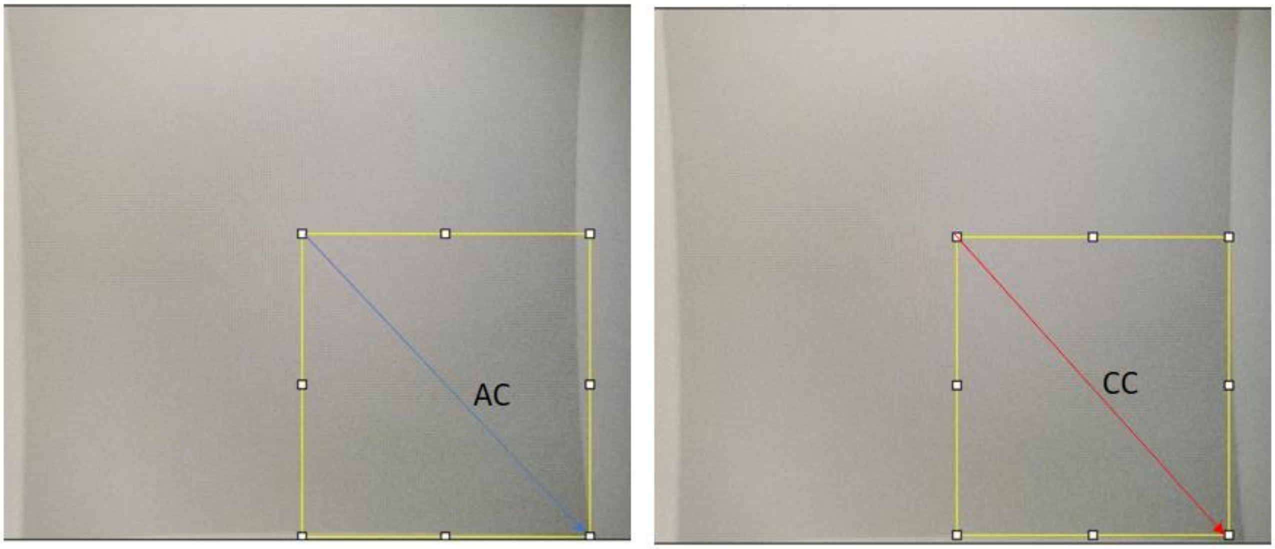

**Figure S8. Quantification of distortion of Mactrem macro lens.** The image of the distortion target was first analyzed via ImageJ to determine distances from the center to the actual corner (AC) of the distortion target image and from the center to the center edge corner (CC). The distortion (D), which is a percentage by convention, is subsequently $D = \frac{AC-CC}{CC} \times 100\%$.

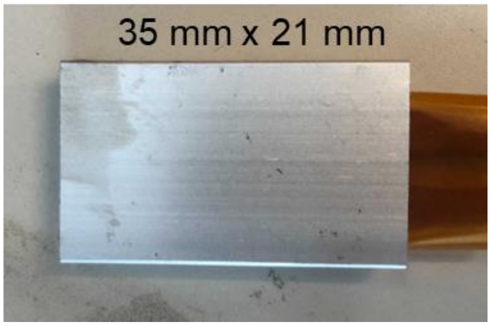

**Figure S9. 50W AC DC 12V PTC ceramic heater.**

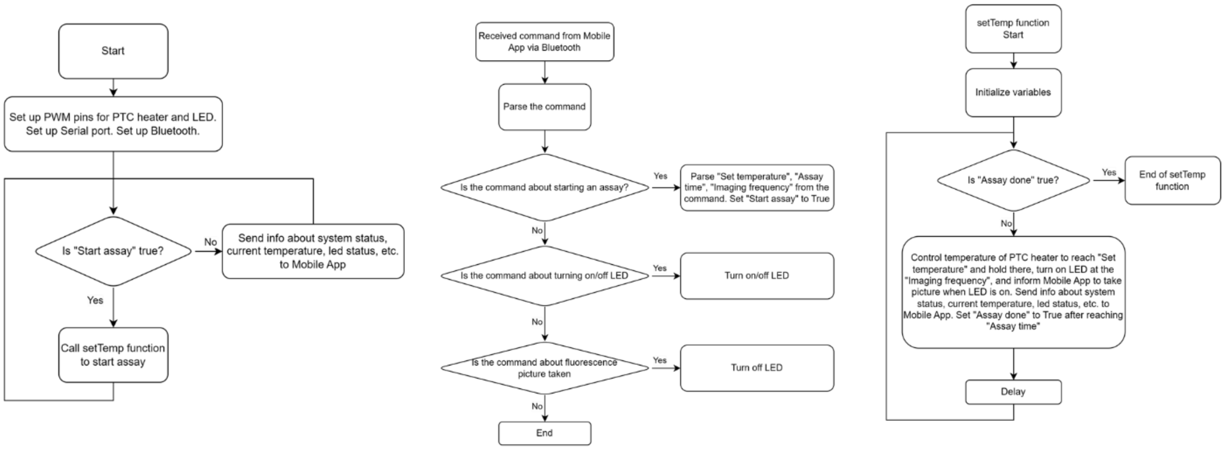

**Figure S10. Flowchart of program on TinyPICO ESP32 that receives commands from smartphone app.** From the received commands, TinyPICO ESP32 controls temperature of PTC ceramic heater for isothermal amplification, turn on/off LED for fluorescence excitation, inform the smartphone app when it is time for image capturing, and report system status to the smartphone app.

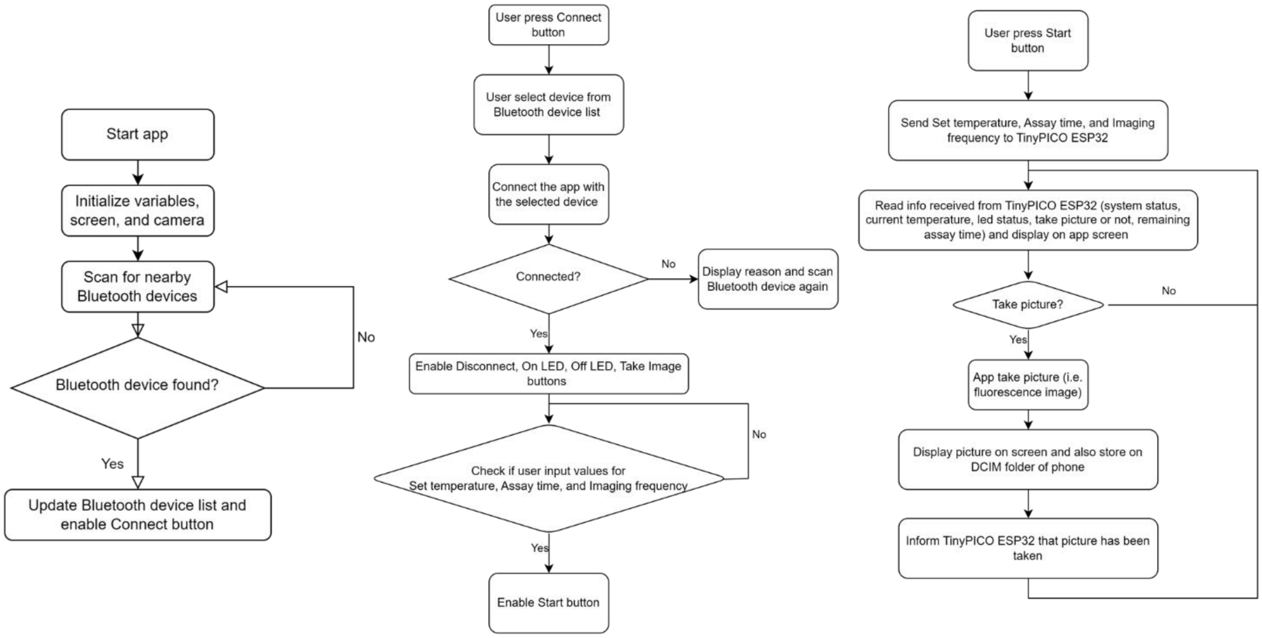

**Figure S11. Flow chart of smartphone app.** The app allows user to set up Bluetooth connection with the TinyPICO ESP32 microcontroller and to set up parameters (temperature, assay time, imaging frequency) for the assay. Once user finish setting the parameters and press “Start”, the app will send these parameters to the TinyPICO ESP32 microcontroller. Based on the received parameters, the TinyPICO ESP32 microcontroller will control temperature and LED (see above). The TinyPICO ESP32 microcontroller will also inform the smartphone app when it is time for image capturing and send system status information to the app for display on app screen.

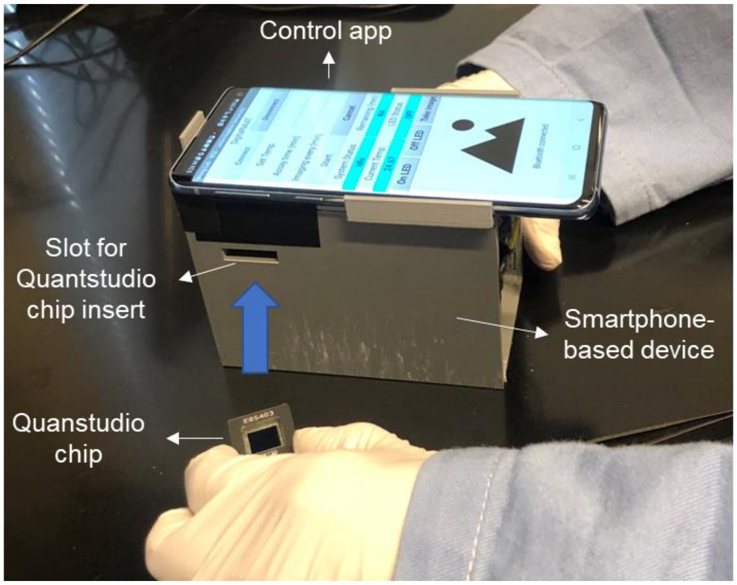

**Figure S12. Smartphone-based device.** The smartphone-based device has one slot on its side for inserting Quantstudio chip loaded with sample. After inserting the chip, user set parameters and start the assay using a customized control app installed on the smartphone.

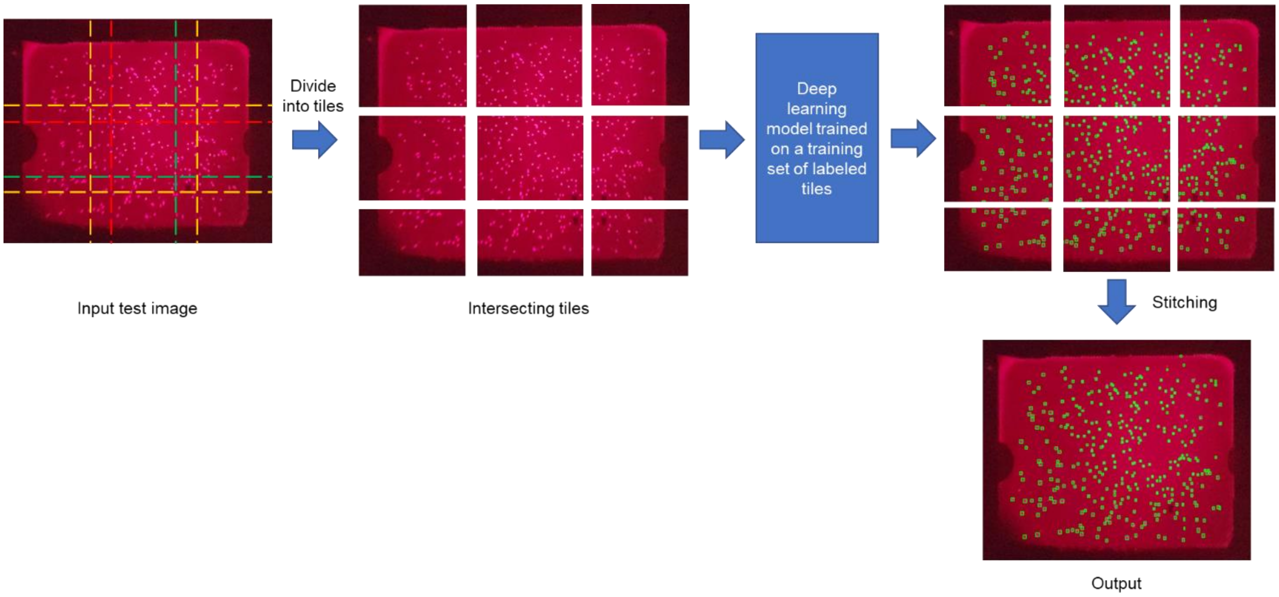

**Figure S13. The concept of small object detection by using deep learning and image tiling.** The input image was divided into small intersecting tiles. The intersecting tiles were then input into a deep learning model that had been trained on a training set of labeled tiles. Outputs of the model were then stitched together to create a final output.

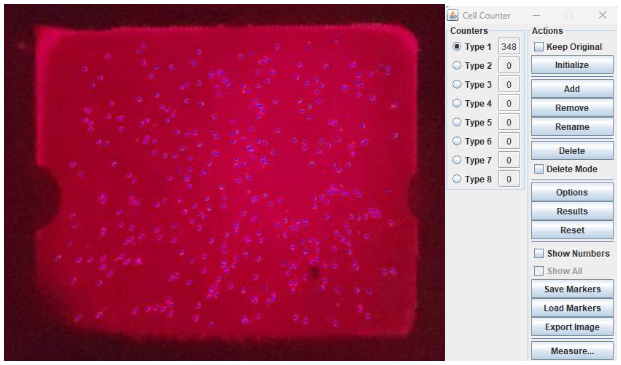

**Figure S14. Manual counting of positive wells using Cell counter tool in ImageJ.**

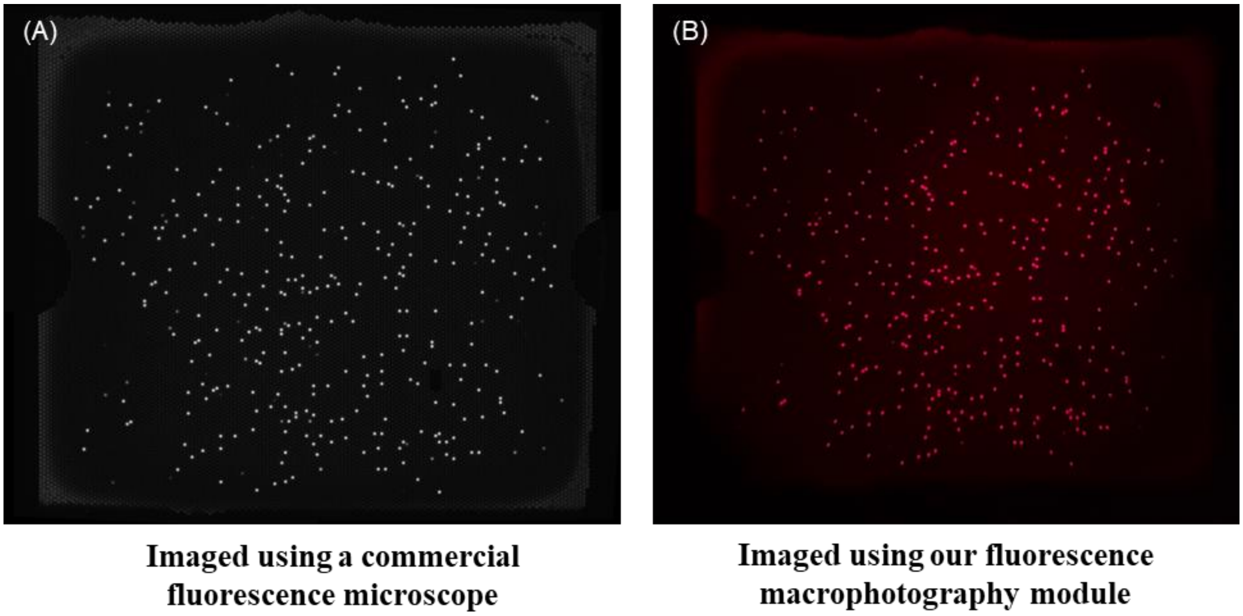

**Figure S15. Fluorescence images of a QuantStudio chip captured by a commercial fluorescence microscope (A) and our fluorescence macrophotography module (B).**

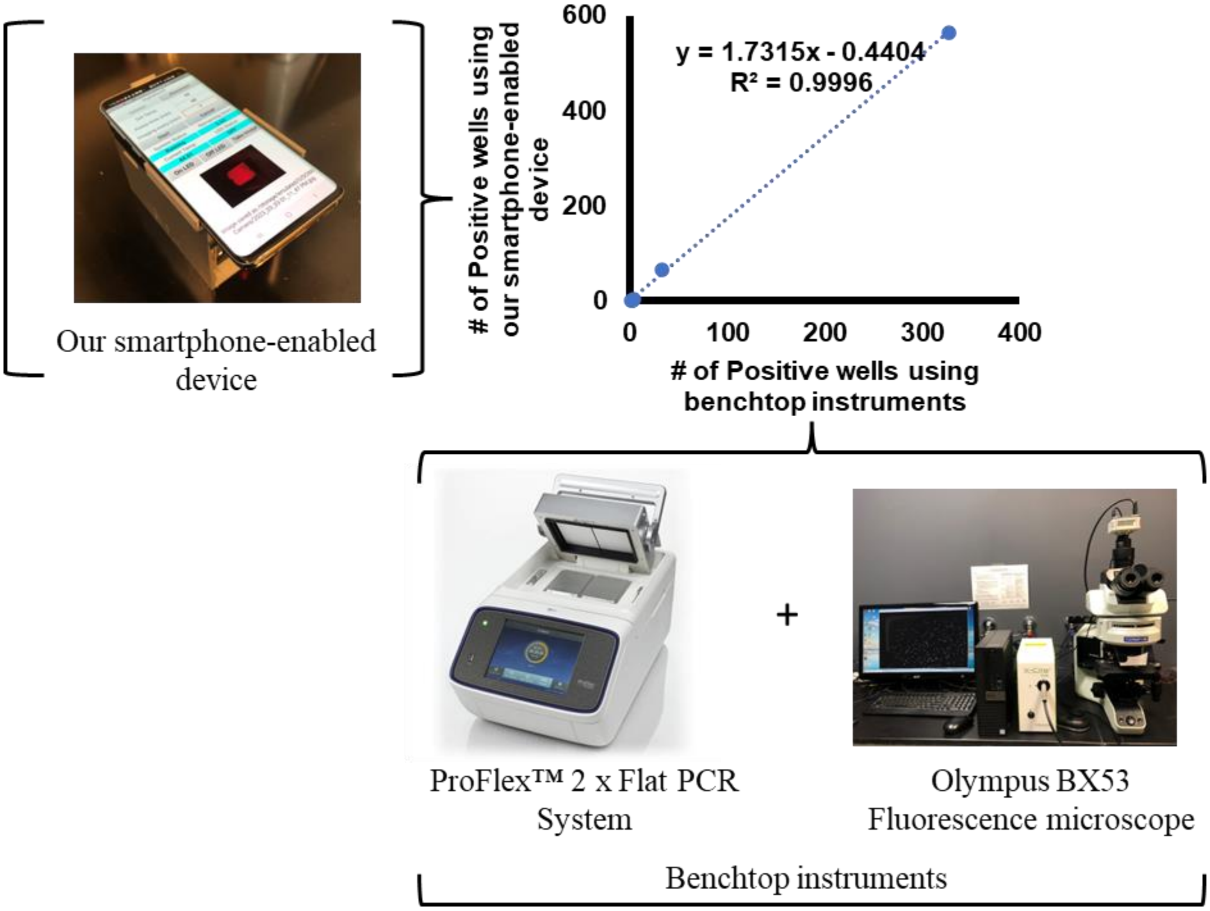

**Figure S16. Comparison between our smartphone-enabled device vs. benchtop instruments.**

### Table S1. Primer and crRNA sequences

| Component | Sequence (5' → 3') | Reference |
|---|---|---|
| RPA Forward Primer | CAAGCAGCCATGCAAATGTTAAAAGAAACCATC | (Ding et al., 2020) |
| RPA Reverse Primer | GTAGTTCCTGCTATGTCACTTCCCCTTGGATC | |
| Cas12a Guide RNA 1 | /AltR1/UAAUUUCUACUAAGUGUAGAU**AUCCCAUUCUGCAGCUUCCUCAUU**/AltR2/ | |
| Cas12a Guide RNA 2 | /AltR1/UAAUUUCUACUAAGUGUAGAU**UUGCACCAGGCCAGAUAAGA**/AltR2/ | |
| Fluorogenic Reporter | /Alex647N/TTATT/IAbRQSp/ | (Hsieh et al., 2020) |
| Fluorogenic Reporter | /Alex647N/TCCCT/IAbRQSp/ | (Sun et al., 2022) |

*Note.* All oligonucleotides synthesized by Integrated DNA Technologies (IDT); Bolded: Target regions in Cas12a guide RNAs; Abbreviations: Cas12a, New England Biolabs' EnGen Lba Cas12a from *Lachnospiraceae bacterium* ND2006; AltR1 and AltR2, IDT's proprietary Alt-R modifications; IAbRQSp, Iowa Black RQ (quencher).

**Table S2. Bill of materials.**

| Component | Product number | Vendor | Unit Cost | Quantity | Subtotal |
|---|---|---|---|---|---|
| **Temperature control** | | | | | |
| PTC heating element 50W AC DC 12V | | Amazon | $12.99 | 1 | $12.99 |
| Thermistor 100K Ohm | GA100K6MCD1 | Mouser | $13.41 | 1 | $13.41 |
| **LED** | | | | | |
| Red (627nm) Rebel LED on a SinkPAD-II 20mm Star Base - 53 lm @ 350mA | SP-01-R5 | Luxeonstar | $7.59 | 1 | $7.59 |
| **Filter cube** | | | | | |
| Fluorescence Dichroic Filters 660nm, 12.5 x 17.6mm | STOCK #67-068 | Edmund Optics | $165.00 | 1 | $165.00 |
| Fluorescence Bandpass Filters 624nm CWL, 12.5mm Dia, 40nm Bandwidth, OD 6 | STOCK #67-021 | Edmund Optics | $215.00 | 1 | $215.00 |
| Fluorescence Bandpass Filters 692nm CWL, 12.5mm Dia, 40nm Bandwidth, OD 6 | STOCK #67-024 | Edmund Optics | $215.00 | 1 | $215.00 |
| **Macro Lens** | | - | | | |
| MACTREM 25× Macro Lens | | Amazon | $29.97 | | $29.97 |
| **Control circuit** | | | | | |
| TinyPICO ESP32 Development Board with USB-C | Product ID: 5028 | Adafruit | $21.95 | 1 | $21.95 |
| IRLB8721PBF MOSFET N-CH 30V 62A TO220AB | IRLB8721PBF | Mouser | $1.08 | 2 | $2.16 |
| BuckPuck 3021-D-E-350, LEDdynamics Inc. | 3021-D-E-350 | Luxeonstar | $10.99 | 1 | $10.99 |
| **Battery** | | | | | |
| Rechargeable 12V 3000mAh Lithium ion Battery Pack | YB1203000-USB | Amazon | $26.99 | 1 | $26.99 |
| **Housing** | | | | | |
| PLA fillament: Prusament PLA Galaxy Silver 1kg | | Prusa | $29.99 | 0.05 | $1.50 |
| | | | | **Total** | $692.58 |
